# Diagnostic Utility of Brain MRI in Spontaneous Intracerebral Hemorrhage: A Retrospective Cohort Study and Meta-Analysis

**DOI:** 10.1101/2023.04.19.23288843

**Authors:** Mitch Wilson, Jia-Yi Wang, Alexander Andreev, Aristeidis H. Katsanos, Magdy Selim, Vasileios-Arsenios Lioutas

## Abstract

**Background:** The diagnostic yield of brain Magnetic Resonance Imaging (MRI) in spontaneous intracerebral hemorrhage (ICH) is unclear. We performed both an independent single-center retrospective cohort study and a meta-analysis to assess the detection rate of secondary lesions on MRI in patients with spontaneous ICH.

**Methods:** In the retrospective cohort study, we examined 856 consecutive patients with spontaneous ICH. Brain MRI scans on admission and follow-up were assessed for secondary lesions. We also examined clinical and CT radiographic variables associated with secondary lesions in univariable analysis. In the meta-analysis we searched PubMed and EMBASE for articles investigating the secondary lesion detection rate on brain MRI in spontaneous ICH. Random-effects models were used to calculate the pooled estimate of secondary lesion detection rate. The meta-analysis used the Preferred Reporting Items for Systematic Reviews and Meta-Analyses (PRISMA) guidelines.

**Results:** Of the 856 patients with ICH, 481 (56%) had at least one brain BRI performed [70±14 years, 270(56% male)]. 462(54%) had an admission MRI and 138(16%) had both admission and follow-up MRIs. The detection rate of secondary lesions on admission MRIs was 24/462(5.2%). 4/127(3.1%) patients with a negative admission MRI had a lesion identified on follow-up MRI. No clinical or radiographic variables were associated with a secondary lesion on MRI using univariable analysis. The meta-analysis included 5 studies total (4 identified in the PubMed and EMBASE searches as well as our own cohort study) comprising 1147 patients with spontaneous ICH who underwent brain MRI. The pooled detection rate of secondary lesions was 11% (95%CI: 7-16%).

**Conclusions:** No predictors of secondary lesion detection were identified in our cohort study. Prospective studies are required to better understand the diagnostic utility of MRI in spontaneous ICH.

## INTRODUCTION

Brain MRI is often performed in patients with spontaneous intracerebral hemorrhage (ICH) to evaluate for an underlying structural etiology such as neoplasm or vascular malformation. Despite the routine use of MRI in spontaneous ICH, and although a recent update of the relevant American Heart Association offers suggestions on the use of MRI in spontaneous ICH (1), there remains knowledge gaps regarding which patients are more likely to benefit from a brain MRI as well as the optimal timing for obtaining imaging.

This ambiguity reflects the paucity of available data, with only a limited number of studies having assessed the ability of MRI to detect secondary lesions (2-5). The majority of these studies were limited by small sample sizes with the exception of one study of 400 patients which demonstrated that MRI/MRA of the brain detected a secondary lesion in 12.5% of cases of spontaneous ICH and led to changes in diagnosis and management in 14% and 20% of cases, respectively (4). Data regarding the utility of follow-up brain MRI is even more limited with only one study to date having assessed this question (6). A meta-analysis examining the diagnostic utility of brain MRI in patients with spontaneous ICH has yet to be performed and is warranted.

The objective of our study is two-fold: 1) to retrospectively assess the rate of detection of secondary lesions in a large cohort of patients with spontaneous ICH on admission and follow-up MRI; and 2) to assess the aggregate rate of detection of secondary lesions on brain MRI in spontaneous ICH using a meta-analysis.

## METHODS

### Retrospective Cohort Study

#### Patients

We included consecutive patients with presumed spontaneous ICH at our tertiary academic hospital between 2008-2018. We excluded patients with known secondary cause of ICH at the time of presentation.

#### Clinical and demographic variables

We collected data on age, sex, comorbidities (hypertension, diabetes, hyperlipidemia, prior history of ischemic stroke or ICH, coronary artery disease), smoking, alcohol or drug abuse, medications (antiplatelets, anticoagulants, antihypertensives and statins), systolic, diastolic and mean blood pressure (SBP, DBP and MBP, respectively) on presentation, Glasgow Coma Scale (GCS) and the ICH score (7).

#### Imaging

##### Hematoma volume and expansion

Hematoma volumes, were computed from the baseline scans and follow-up scans performed within 72 h from the baseline scan using the ABC/2 method (8). The interrater agreement in twenty randomly selected CT scans was 0.97.

#### Hematoma location

Location was recorded as deep (involving the basal ganglia, thalamus, internal capsule, corona radiata), lobar (cortex and juxtacortical areas) or infratentorial.

#### MRI time, acquisition and analysis

All patients underwent MRI according to a standardized protocol as part of routine clinical assessments. The protocol included T1- and T2-weighted, 3D T1 Magnetization Prepared - RApid Gradient Echo (MP-RAGE) sequences and fluid-attenuated inversion recovery (FLAIR), gradient-echo T2*-weighted (GRE or SWI), axial trace DWI with 2 b values (0 and 1000), and apparent diffusion coefficient (ADC) sequences. Studies were performed on 1.5 or 3T scanners. Sequences typically included 24–30 slices of 5-mm thickness with a matrix size of 128 × 128.

The imaging parameters were as follows: T1 [repetition time (TR) 420 ms; echo time (TE) 8.8 ms]; T2 (TR 4500 ms; TE 95 ms); FLAIR (TR 9000 ms; TE 84 ms) GRE (TR 835 ms; TE 26 ms); diffusion tensor imaging (TR4528 ms; TE 103 ms). All patients received intravenous gadolinium unless a contraindication (allergic reaction or impaired renal function) were present.

#### Secondary lesion assessment

Among patients with spontaneous ICH who underwent a brain MRI, the radiology reports were reviewed to evaluate for the presence of a secondary lesion on baseline and follow-up MRI. Secondary lesions were considered those that had a plausible causative link with the index ICH and included brain tumors, ischemic strokes with hemorrhagic transformation, cerebral venous sinus thrombosis, and underlying vascular malformations.

#### Outcomes of interest

Our main outcome was the rate of detection of secondary lesion on admission MRI and the rate of detection of secondary lesion on follow-up MRI for patients with a non-diagnostic admission MRI. As a secondary outcome we assessed for baseline clinical and radiographic predictors of secondary lesion on MRI.

#### Statistical Analysis

Categorical and ordinal variables are presented as percentages and continuous variables as either mean ± standard deviation (SD) or median and interquartile range (IQR) depending on normality of distribution. We examined univariable associations between baseline clinical, imaging and demographic variables and our outcomes of interest to determine if clinical or CT radiographic variables were associated with the presence of a secondary lesion. For continuous variables, we used Student’s t-test or Wilcoxon rank sum test; for categorical variables we used the chi-squared test. Statistical significance was set at a two-sided p value level of 0.05. Multivariate logistic regression was performed for variables with a p value of <0.1 in univariable analysis. Analyses were performed using Stata 13 (StataCorp, College Station, TX, USA).

The study was approved by the institutional review board of Beth Israel Deaconess Medical Center (BIDMC).

### Meta-Analysis

We systematically searched for studies that have assessed the detection rate of secondary lesions on brain MRI in patients with spontaneous ICH. The meta-analysis was reported according to the PRISMA guidelines (9).

#### Inclusion Criteria

Studies were included if they: 1) enrolled patients with spontaneous non-traumatic ICH and had a brain MRI performed; 2) reported the prevalence of secondary lesions on MRI; and 3) were primary studies that did not use duplicate or pooled data from a shared cohort.

#### Search Strategy

We searched both PubMed and EMBASE on January 26, 2023 using the following MESH terms (PubMed)/keywords (EMBASE): *(intracerebral hemorrhage OR intracranial hemorrhage OR ICH OR intracranial bleeding OR intracranial bleed) AND (MRI OR magnetic resonance imaging) AND (utility OR yield)*.

#### Data Extraction

The abstracts of all records were reviewed by one author; all studies that were not relevant to the utility of brain MRI in spontaneous ICH were excluded. The full texts of the remaining studies were reviewed by two authors (MW and VL) to determine eligibility for the meta-analysis (see Figure 1). For all included studies we recorded the prevalence of secondary lesions (our primary outcome) as well as baseline cohort characteristics and any predictors of secondary lesion detection, if available. A quality assessment was made for all included studies using the Newcastle-Ottawa Scale (NOS) adapted for cross-sectional studies (10). Studies with NOS scores of 0–4, 5–6, 7–8, and 9-10 were considered as unsatisfactory, satisfactory, good or very good quality, respectively.

**Figure 1.**
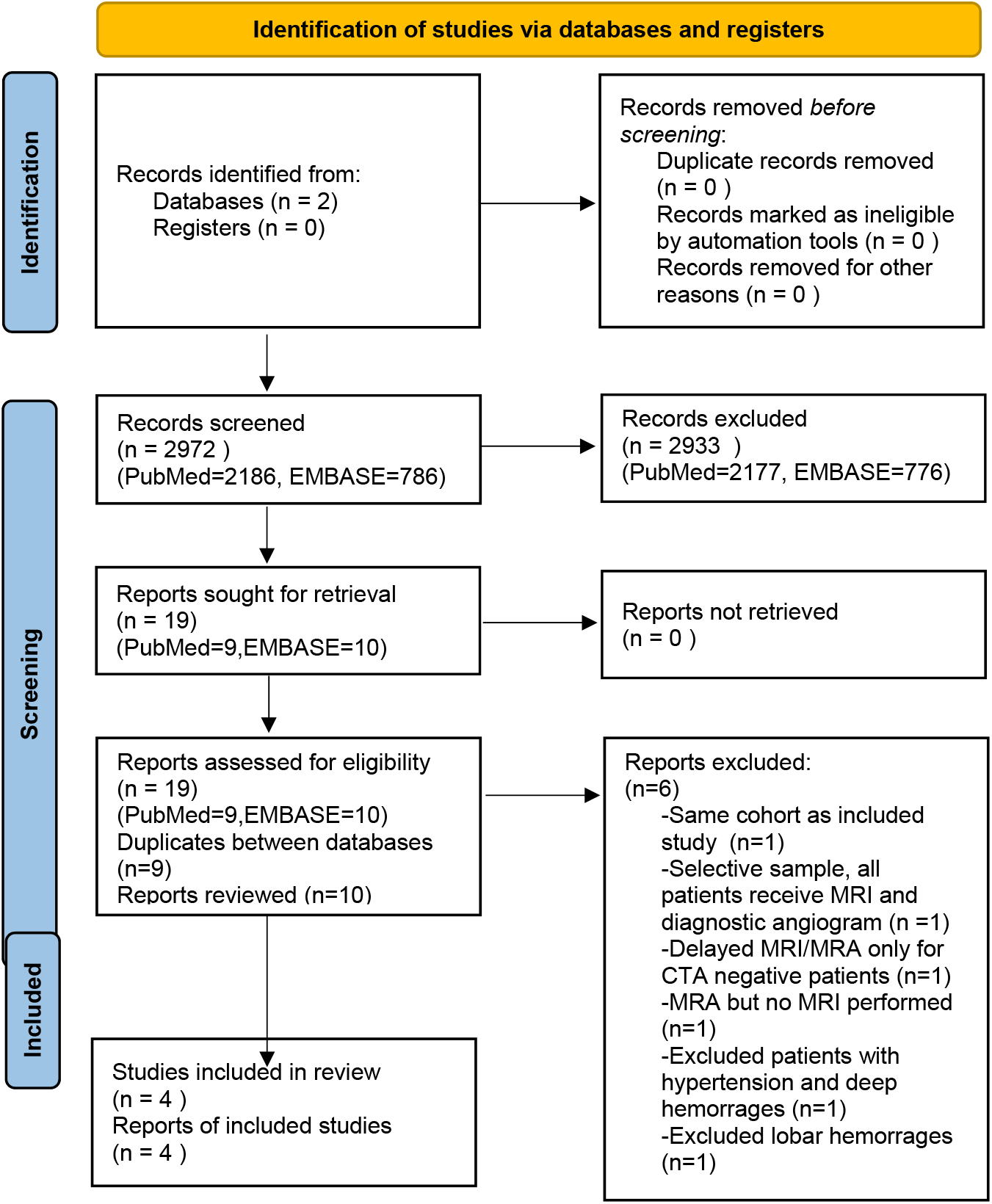
PRISMA Flowsheet describing process for study selection for meta-analysis.

#### Statistical Analysis

We calculated the detection rates of secondary lesions on MRI for each individual study by dividing the number of patients with any detectable lesion on MRI by the total number of patients receiving MRI after a spontaneous ICH. For all proportion analyses we implemented the variance-stabilizing double arcsine transformation (11). The random-effects model

(DerSimonian and Laird) was used to calculate the pooled estimates in both the overall and subgroup analyses. We assessed heterogeneity between studies with the Cochrane Q and I2 statistics.

## RESULTS

### Retrospective Cohort Study

Our cohort comprised 856 spontaneous ICH patients diagnosed on CT head imaging. Among these, 481(56%) had at least one brain MRI [70±14 years, 270(56% male)]: 462 (54%) at baseline, 156(18%) on follow-up and 138(16%) on both. Baseline clinical and radiographic variables for all patients with at least one MRI are presented in Table 1.

**Table 1:** Baseline clinical and radiographic variables for all patients with spontaneous ICH who underwent brain MRI.

|  | Full cohort<br>characteristics<br>(n=481) | Abnormality<br>(n=28) | No<br>abnormality<br>(n=453) | p value |
| --- | --- | --- | --- | --- |
| Age, mean (SD) | 69.5 (14.2) | 65.7 (13.8) | 69.7 (14.2) | 0.15 |
| Female sex, n (%) | 211 (44) | 9 (32) | 202 (45) | 0.24 |
| Hypertension, n (%) | 349 (73) | 22 (79) | 327 (72) | 0.52 |
| Diabetes, n(%) | 107 (22) | 4 (14) | 103 (23) | 0.34 |
| Baseline hematoma volume (ml),<br>median (IQR) | 12.7 (4.8-35.7) | 8.11 (4.2-23.4) | 12.9 (4.9-36) | 0.36 |
| Follow up hematoma volume (ml),<br>median (IQR) | 15.7 (5.4-40.8) | 10.4 (5.9-19.6) | 16.5 (5.39-42.1) | 0.24 |
| Hematoma growth>12.5ml, n(%) | 52 (12) | 3 (15) | 49 (12) | 0.72 |
| Hematoma growth >6ml, n(%) | 79 (19) | 3(15) | 76 (19) | 1 |
| Hematoma growth >33%, n(%) | 96 (22) | 7 (33) | 89 (22) | 0.28 |
| ICH score, median (IQR) | 1 (0-2) | 1 (0-2) | 1 (0-2) | 0.38 |
| GCS, median (IQR) | 15 (13-15) | 15 (13-15) | 15 (13-15) | 0.99 |
| Lobar, n(%) | 255 (53) | 16 (57) | 239 (53) | 0.7 |
| IVH, n(%) | 134 (26) | 7 (25) | 127 (28) | 0.83 |
| Infratentorialn (%) | 49 (11) | 5 (18) | 46 (10) | 0.21 |

Characteristics of patients who received MRI vs those who did not are presented in Table 2. Patients who underwent MRI were more likely to be younger, have absence of hypertension, have smaller hemorrhage size, lower ICH score, higher GCS, and have lobar location of hemorrhage. Table 3 compares patients with lobar ICH who underwent brain MRI to those who did not. Patients with lobar ICH who underwent brain MRI were more likely to be younger, have smaller baseline and follow up hematoma volume, less hematoma expansion, lower ICH score, and higher GCS than patients with lobar ICH who did not receive an MRI.

**Table 2:**
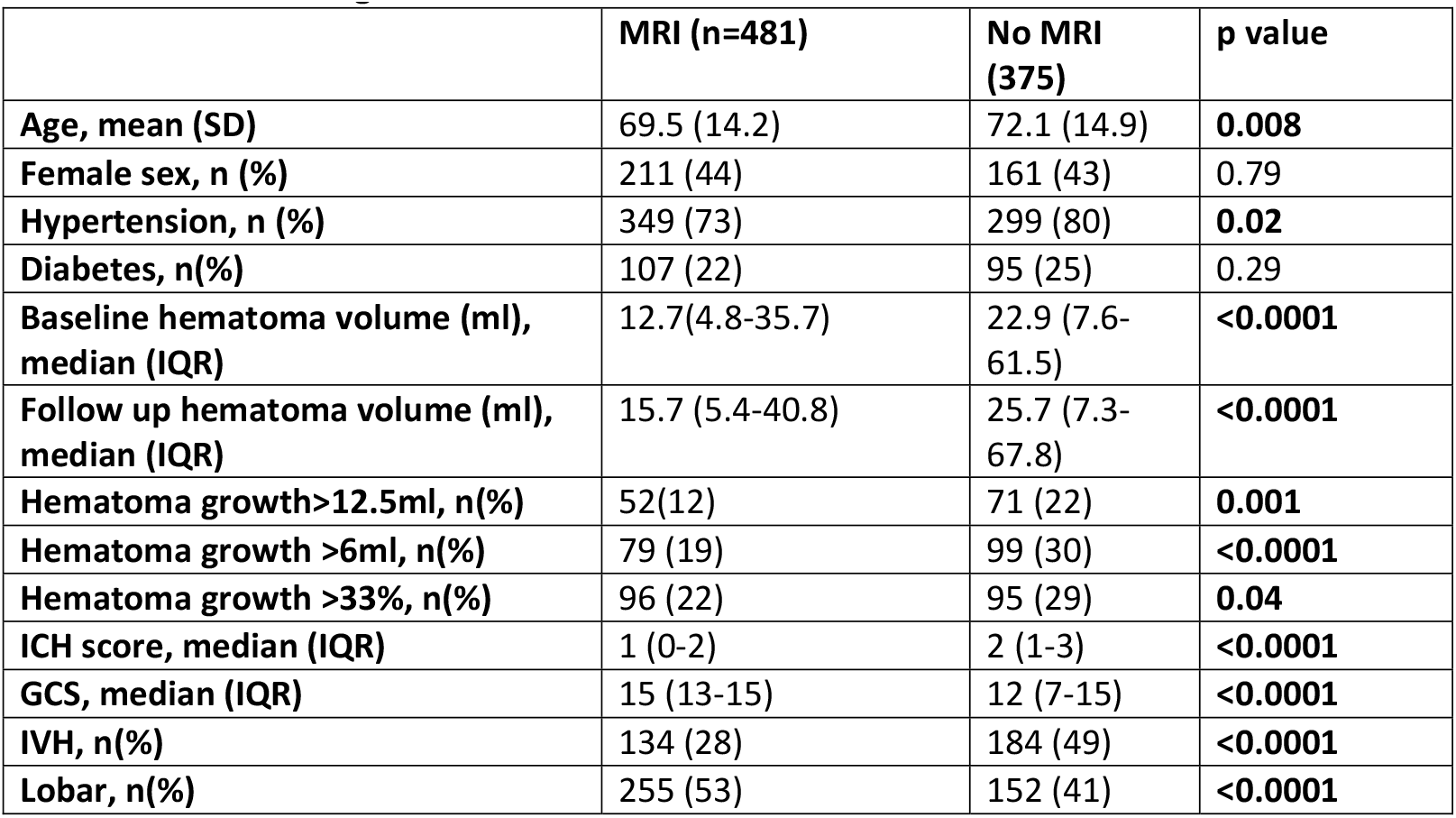
Baseline clinical and radiographic variables for patients with spontaneous ICH who did and did not undergo MRI of the brain.

**Table 3:** Baseline clinical and radiographic variables for patients with spontaneous lobar ICH who did and did not undergo MRI of the brain.

|  | **Lobar ICH all (n=407)** | **Lobar with MRI** | **Lobar without MRI** | **p value** |
| --- | --- | --- | --- | --- |
| **Age, mean (SD)** | 73.5 (14.2) | 71.8 (13.3) | 76.4 (15.1) | **0.002** |
| **Female sex, n (%)** | 201 (49) | 119 (47) | 82 (54) | 0.18 |
| **Hypertension, n (%)** | 286 (70) | 175 (69) | 111 (73) | 0.37 |
| **Diabetes, n(%)** | 89 (22) | 52 (20) | 37 (24) | 0.39 |
| **Baseline hematoma volume (ml), median (IQR)** | 32.3 (11.6-66.3) | 23.0 (9.5-51.2) | 58.9 (24.9-112.2) | **<0.0001** |
| **Follow up hematoma volume (ml), median (IQR)** | 35.6 (13.1-69.4) | 27.6 (10.3-54.2) | 66.6 (22.2-112.1) | **<0.0001** |
| **Hematoma growth>12.5ml, n(%)** | 80 (23) | 40 (18) | 40 (30) | **0.01** |
| **Hematoma growth >6ml, n(%)** | 109 (31) | 57 (26) | 52 (39) | **0.009** |
| **Hematoma growth >33%, n(%)** | 81 (23) | 47 (21) | 34 (26) | 0.36 |
| **ICH score, median (IQR)** | 2 (1-3) | 1 (0-2) | 2.5 (2-3) | **<0.0001** |
| **GCS, median (IQR)** | 14 (9-15) | 14.5 (12-15) | 10 (7-14) | **<0.0001** |
| **IVH, n(%)** | 122 (30) | 57 (22) | 65 (43) | **<0.0001** |

24/462 (5.2%) of baseline MRIs revealed a secondary lesion. Secondary lesions included mass in 6 (1.4%) patients, cavernoma in 8 (1.9%) patients, arteriovenous malformation in 6 (1.4%) patients, hemorrhagic transformation of ischemic stroke in 3 (0.7%) patients, and Moyamoya disease in 1 (0.2%) patient.

Among the 138 patients with both baseline and follow-up MRIs, 127 had a negative admission MRI; 4 (3.1%) of those patients with a negative MRI at baseline had a lesion identified on follow-up MR. Secondary lesions detected in follow up MRI included mass (3/127, 2.4%) and cavernoma (1/127, 0.8%).

For the 481 patients who underwent MRI, no baseline clinical or radiographic variables were associated with the presence of a secondary lesion in univariable analysis (table 1).

### Meta-Analysis

#### Outcome analysis

The PubMed and EMBASE literature searches identified 4 cohort studies meeting our inclusion criteria that assessed the prevalence of secondary lesions on brain MRI in spontaneous ICH (Figure 1). We also included our own cohort study and therefore the meta-analysis consisted of a total of 5 studies. All 5 studies had NOS scores of 8/10 and so were deemed to be of good quality. Each study lost 2 points in the selection category – one point for not having a justified sample size and one point for not having a description of the response rate or the characteristics of the responders and non-responders. There was a total of 1147 patients with spontaneous ICH who underwent a baseline brain MRI. The rates of detection for each of the 5 retrospective studies is provided in table 4 along with baseline demographic variables. A forest plot is provided in figure 2. The pooled detection rate of secondary lesions on brain MRI in patients with spontaneous ICH was 11.0% (95%CI: 7-16%). There was presence of between-study heterogeneity in the rates of structural lesion detection (I^2^= 82%, p=0.00).

**Table 4.**
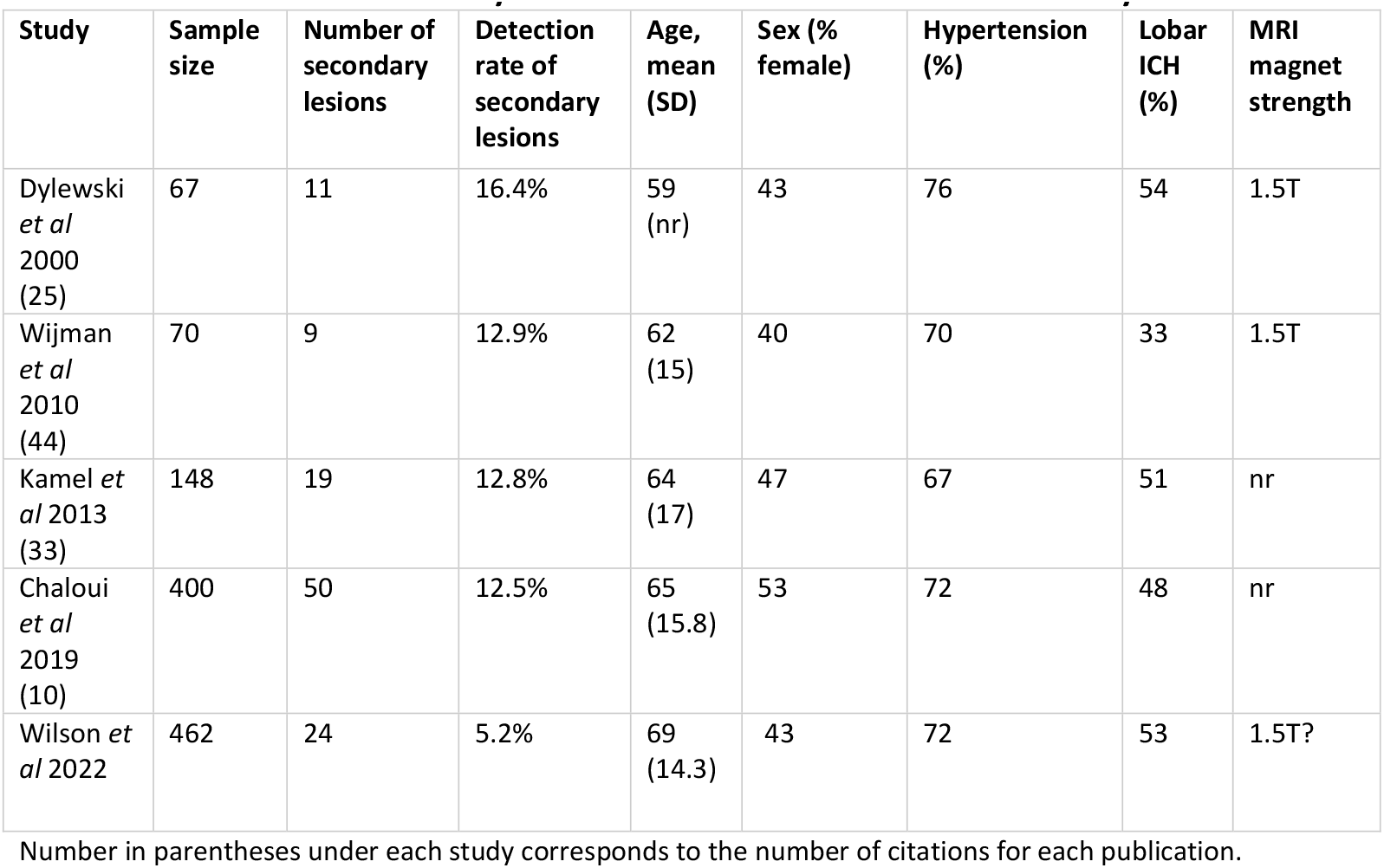
Prevalence of secondary lesions in 5 studies included in meta-analysis.

| Study | Sample size | Number of secondary lesions | Detection rate of secondary lesions | Age, mean (SD) | Sex (% female) | Hypertension (%) | Lobar ICH (%) | MRI magnet strength |
| --- | --- | --- | --- | --- | --- | --- | --- | --- |
| Dylewski *et al* 2000 (25) | 67 | 11 | 16.4% | 59 (nr) | 43 | 76 | 54 | 1.5T |
| Wijman *et al* 2010 (44) | 70 | 9 | 12.9% | 62 (15) | 40 | 70 | 33 | 1.5T |
| Kamel *et al* 2013 (33) | 148 | 19 | 12.8% | 64 (17) | 47 | 67 | 51 | nr |
| Chaloui *et al* 2019 (10) | 400 | 50 | 12.5% | 65 (15.8) | 53 | 72 | 48 | nr |
| Wilson *et al* 2022 | 462 | 24 | 5.2% | 69 (14.3) | 43 | 72 | 53 | 1.5T? |

Number in parentheses under each study corresponds to the number of citations for each publication.

**Figure 2.**
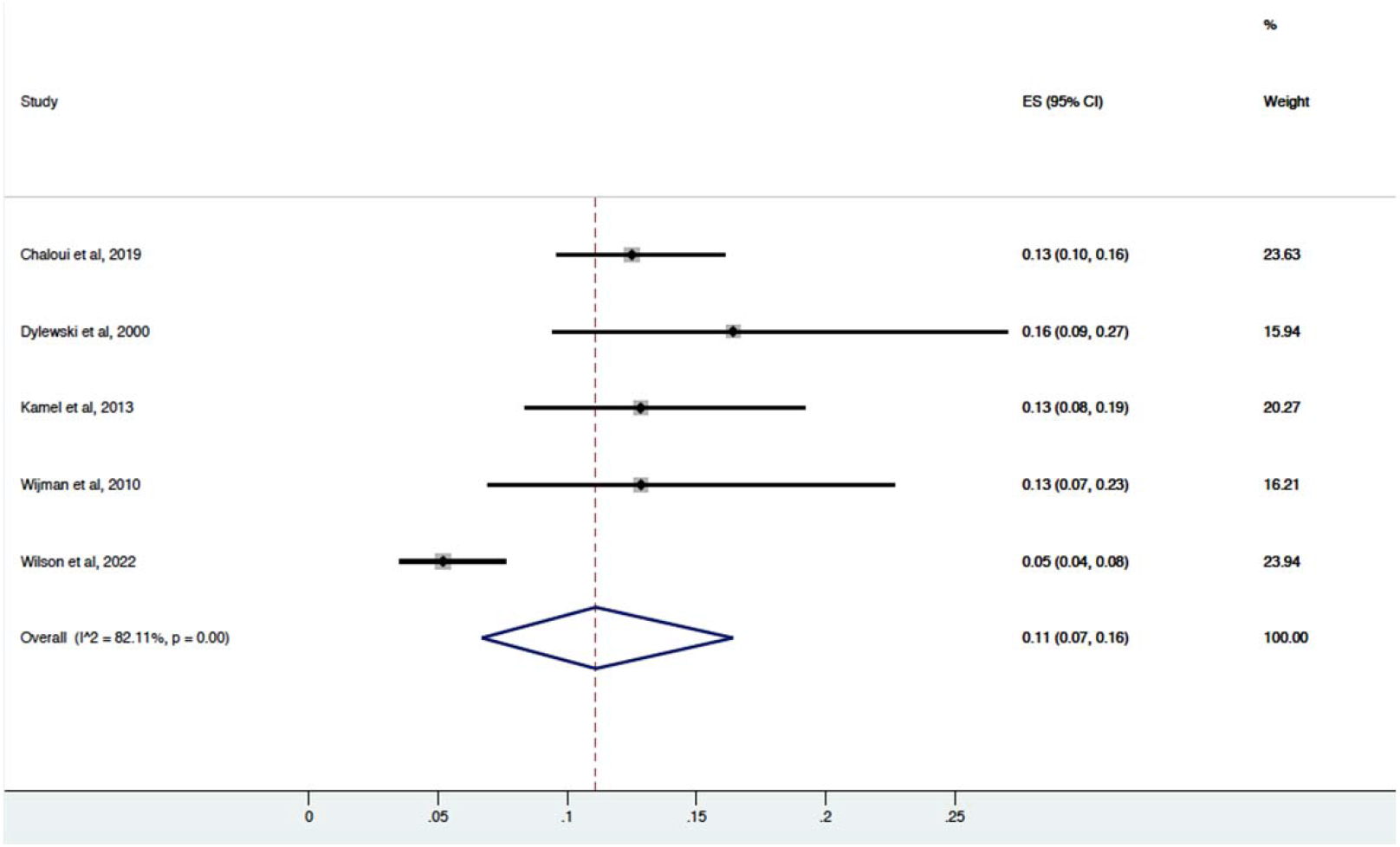
Forest Plot demonstrating effect size and weight of the 5 studies included in the meta-analysis.

## DISCUSSION

In this retrospective single-center cohort of 481 patients with spontaneous ICH who underwent brain MRI we found that the rate of secondary lesion detection was 5.2% on admission MRI and 3.1% on follow-up MRI for patients who had a negative admission MRI. No baseline clinical or radiographic predictors of secondary lesion on MRI were identified in the cohort. A meta-analysis of 5 retrospective cohort studies comprising 1147 patients recorded a pooled rate of secondary lesion detection on brain MRI performed during admission for patients with spontaneous ICH to be 11.0%.

The clinical significance of detecting a secondary lesion in 11% of patients with spontaneous ICH who undergo brain MRI and whether it justifies a universal brain MRI is a complex issue. Some could argue that the cost of obtaining 100 MRIs to detect 11 lesions is justified if the detection of these lesions truly impacts patient outcomes. However, others may argue that a detection rate of 11% is too low to justify the cost of a universal brain MRI strategy in ICH. Our meta-analysis and retrospective cohort study did not assess whether MRI affected patient management or outcome. A single small study of 70 patients included in our meta-analysis found that MRI changed patient management in 20% of cases (3) but did not specifically assess the impact of detection of secondary lesions on long-term patient outcomes. No other studies have assessed the effect of secondary lesion detection on patient management and long-term outcomes or evaluated the cost-effectiveness of MRIs for patients with spontaneous ICH. Additional studies are needed to better understand the utility of brain MRI in these patients.

No baseline predictors of secondary lesion detection on MRI were identified in our retrospective cohort study. We therefore cannot provide specific guidance on when an MRI should or should not be performed in patients with spontaneous ICH or which patients are more likely to have a secondary lesion detected. Our results contrast with those of Chalouhi *et al*. who found that younger age, absence of hypertension, and non-basal ganglia/thalamic location of hemorrhage were predictive of secondary lesion detection (4). The discrepancy between our results might be due to the fact that our cohort was selective, having already excluded patients with known secondary cause, while favoring lobar ICH. Conversely, in the study by Chalouhi *et al*. MRI was obtained for all patients with spontaneous ICH as part of an institutional protocol which may have contributed to the discrepant findings between our studies. It is plausible that a universal MRI policy in our institution might have reproduced the finding that lobar ICH is a more likely harbinger of a secondary lesion. The authors recommended against the use of routine MRI in patients with basal ganglia/thalamic hemorrhages over 65 years of age and in all patients over 85 years of age. as no secondary lesions were identified in these subsets of patients. Our cohort study, however, found that admission MRI detected a secondary lesion in 2 patients over the age of 65 with non-lobar hemorrhage and in 4 patients over the age of 85.

We found that the rate of lesion detection on follow-up MRI for patients with an initial non-diagnostic MRI (3.1%) was lower than the rate of lesion detection on initial admission MRI (5.2%). These findings are similar to those of Mouchtouris *et al*. who found that of 113 patients with a non-diagnostic admission MRI no patients (0%) had a secondary lesion identified on follow-up MRI (6). Similar to the diagnostic utility of an admission MRI in patients with spontaneous ICH, current data do not provide decisive support either for or against a universal policy of follow-up MRI for these patients. The relatively low yield of follow-up MRI in our study and the study by Mouchtouris *et al*and should encourage clinicians to use clinical judgement when deciding whether a follow-up MRI is appropriate. Further studies with larger number of patients with follow-up MRI are required to better understand the true diagnostic utility of this imaging strategy, its impact on patient outcomes and its cost-effectiveness.

The main limitation of our cohort study was the potential of selection bias due to the lack of standardized criteria for obtaining MRI in ICH patients at our institution. The retrospective nature of our cohort study did not provide measures of patient outcomes and we did not assess the cost-effectiveness of brain MRI in patients with spontaneous ICH. The small number of patients with a follow-up MRI hindered the identification of baseline predictors of secondary lesion detection on follow-up MRI. We did not measure the prevalence of cerebral microbleeds or markers of small vessel disease on MRI which are becoming increasingly relevant in the diagnosis and prognosis of cerebral amyloid angiopathy. (12). Thus, the diagnostic and prognostic relevance of brain MRI in patients with spontaneous ICH will likely extend beyond the identification of a secondary lesion. The meta-analysis had a limited number of included studies and only two studies reported baseline predictors of secondary lesion detection on MRI, precluding data pooling for baseline characteristics predictor analysis. The study by Mouchtouris et al. was the only study with institutional criteria for obtaining MRI in ICH patients, and therefore, other included studies may also have selection bias.

In conclusion, our meta-analysis of 5 retrospective cohort studies found that the rate of detection of secondary lesions on brain MRI in patients with spontaneous ICH is approximately 11%. Detection of a secondary lesion is more likely on the admission MRI and a relatively small number of lesions are identified in follow up MRIs when the baseline scan is negative. We did not identify any baseline clinical or radiographic baseline predictors of lesion detection on MRI. Larger studies assessing long-term outcomes with cost-effectiveness analysis are required to better understand the diagnostic utility of brain MRI in patients with spontaneous ICH.

## Data Availability

The data that support the findings of this study are openly available in Harvard Dataverse. Link URL provided below.

https://dataverse.harvard.edu/dataset.xhtml?persistentId=doi%3A10.7910%2FDVN%2FLIUVMZ&version=DRAFT

## References

1. Greenberg SM, Ziai WC, Cordonnier C, Dowlatshahi D, Francis B, Goldstein JN, et al. 2022 Guideline for the Management of Patients With Spontaneous Intracerebral Hemorrhage: A Guideline From the American Heart Association/American Stroke Association. Stroke. 10.1161/STR.0000000000000407.

2. Kamel H, Navi BB, Hemphill JC. A rule to identify patients who require magnetic resonance imaging after intracerebral hemorrhage. Neurocritical care. 2013;18(1):59–63.

3. Wijman CA, Venkatasubramanian C, Bruins S, Fischbein N, Schwartz N. Utility of early MRI in the diagnosis and management of acute spontaneous intracerebral hemorrhage. Cerebrovascular Diseases. 2010;30(5):456–63.

4. Chalouhi N, Mouchtouris N, Al Saiegh F, Das S, Sweid A, Flanders AE, et al. Analysis of the utility of early MRI/MRA in 400 patients with spontaneous intracerebral hemorrhage. Journal of neurosurgery. 2019;132(6):1865–71.

5. Dylewski DA, Demchuk AM, Morgenstern LB. Utility of magnetic resonance imaging in acute intracerebral hemorrhage. Journal of Neuroimaging. 2000;10(2):78–83.

6. Mouchtouris N, Saiegh FA, Chalouhi N, Sweid A, Papai EJ, Wong D, et al. Low diagnostic yield in follow-up MR imaging in patients with spontaneous intracerebral hemorrhage with a negative initial MRI. Neuroradiology. 2021;63(7):1009–12.

7. Hemphill III JC, Bonovich DC, Besmertis L, Manley GT, Johnston SC. The ICH score: a simple, reliable grading scale for intracerebral hemorrhage. Stroke. 2001;32(4):891–7.

8. Kothari RU, Brott T, Broderick JP, Barsan WG, Sauerbeck LR, Zuccarello M, et al. The ABCs of measuring intracerebral hemorrhage volumes. Stroke. 1996;27(8):1304–5.

9. Page MJ, McKenzie JE, Bossuyt PM, Boutron I, Hoffmann TC, Mulrow CD, et al. The PRISMA 2020 statement: an updated guideline for reporting systematic reviews. Systematic reviews. 2021;10(1):1–11.

10. Wells GA, Shea B, O’Connell D, Peterson J, Welch V, Losos M, et al. The Newcastle-Ottawa Scale (NOS) for assessing the quality of nonrandomised studies in meta-analyses. Oxford; 2000.

11. Freeman MF, Tukey JW. Transformations related to the angular and the square root. The Annals of Mathematical Statistics. 1950:607–11.

12. Charidimou A, Boulouis G, Frosch MP, Baron J-C, Pasi M, Albucher JF, et al. The Boston criteria version 2.0 for cerebral amyloid angiopathy: a multicentre, retrospective, MRI– neuropathology diagnostic accuracy study. The Lancet Neurology. 2022;21(8):714–25.

